# Public Awareness and Attitude towards Counterfeit Medicines in Sudan: A cross-sectional study

**DOI:** 10.1101/2020.09.28.20201400

**Authors:** Wala W. Wagiella, Shaza W. Shantier, Elrasheed A. Gadkariem

**Affiliations:** Department of Pharmacy Practice, Faculty of Pharmacy, University of Khartoum, Sudan; Department of Pharmaceutical Chemistry, Faculty of Pharmacy, University of Khartoum, Sudan

**Keywords:** Counterfeit medicines, Public, Awareness, Attitude, Sudan

## Abstract

**Background:** Counterfeit medicines (CFMs) are a global problem with significant and well-documented consequences for global health and patient safety, including drug resistance and patient deaths. Reports of counterfeit pharmaceuticals in Africa indicate a wide variety of detrimental effects.

**Objectives:** The aim of the present study was to assess the extent, awareness and attitude of public in Sudan towards CFMs.

**Methods:** A cross-sectional study was conducted applying pretested and structured questionnaire. The awareness and attitude were assessed statistically and the association between those and different demographic characteristics was calculated using Fisher exact test and Spearman correlation.

**Results:** A total of 386 participants have enrolled in the study. The majority of the respondents (58%) were found to be aware about the term CFMs with social media mentioned to be the main source. 73% of the respondents considered CFMs of worse quality suggesting getting the medicine from a trustworthy pharmacist in order to avoid buying CFMs. 56% reported their ability to distinguish CFMs from the side effects rather than the package and cost. Education was suggested by 82% of the respondents to have vital role in combating CFMs spread through workshops and campaigns. 68% of the participants were found to have a fair awareness about CFMs. Furthermore, 80% of them showed a good attitude toward CFMs

**Conclusion:** Current literature includes gaps in knowledge and attitude towards CFMs. Therefore attention and concentrated efforts are required on the part of the government, drug manufacturers and health care providers’ especially pharmaceutical analysts to ensure that only drugs of acceptable quality reach the patient.

## Introduction

Counterfeit medicines (CFMs) are of lower quality than their originals, fraudulently manufactured with fake packaging and usually no or a wrong active pharmaceutical ingredient [1]. They can be generic and branded products and according to World Health Organization (WHO) there are six categories including product without active ingredients 32%, or with wrong ingredients 21.4 %, without active ingredients, with insufficient active ingredients 20.2% or with fake packaging 15.6%, products with a high level of impurities or contaminants 8.5% and copies of original product 1%. Counterfeiting is a substantial problem that is growing worldwide, and affects both developed and less developed countries [2]. Recently the trade in counterfeiting has developed into an extensive threat to the public health and the pharmaceutical industry [3]. WHO estimated that the global trade in CFMs is experiencing continuous growth [4], up to 10% of the drugs worldwide may be counterfeits [5, 6]; 50% of them involved antimicrobial drugs, and 78% were from developing countries. Moreover, 59% of cases with available information on the quality of drugs were fraudulent, and only7% had the standard concentration of the active drug [7–9]. However, reporting of counterfeit drugs within WHO is only 15% [10, 11]. The European Commission estimated that counterfeiting in general represents around 5–7% of world trade and around 15% of the global medicines supply chain could be counterfeit [12].

Literature review was conducted to determine the prevalence of CFMs in the United States of America (USA), Europe, Africa and Asia with a focus on the developing countries [13]. The research showed CFM are available worldwide and are most prevalent in developing countries due to weak medicine regulation, control and enforcement. A study was carried out in Saudi Arabia indicated that 34.4% of drugs were counterfeits in 2005 and increase in 2006 to 49.3%. In Africa and Asia the figure exceeds 50% trading in counterfeit and/or substandard drugs annual earnings reached over 35 billion dollars [13, 14].

Advanced computer technologies and the widespread use of the internet lead to the rapid increase of CFM which result in regulatory systems weakness and lack of awareness among health workers and public [15–19]. Unfortunately, the illegal trade in counterfeits has now extended to herbal drugs which are mostly used in developing countries [20].

A study was carried out to assess the prevalence of counterfeit antimalarial medicines where malaria is endemic in 106 countries. In Southeast Asia, it was found that 53% of 188 tested Artesunate tablets were counterfeits (didn’t contain any active ingredient) and 9% of 44 Mefloquine tablets were counterfeits (contained less than 10% of active ingredient) [21]. In sub-Saharan Africa, 35% of 2297 tested samples had failed chemical analysis, 36% of 77 failed packaging analysis, and 20% of 389 were classified as falsified [22].

The primary problem with CFM is the significant health threat they pose and it is not confined to the individual patient, but extends to the whole society, endangering public health and safety [23]. The consequences of using CFM may range from treatment failure, prolonged illness, unusual side effects, anti-microbial/antibiotic resistance leading to mortality from the disease or the toxic components of the CFM [20, 23–26]. People with desperate needs, who cannot afford the full price, tend to be the most vulnerable victims [27]. Reports of CFM in Africa show a wide variety of negative impacts. In addition to those in public health, these incorporate lost taxes to governments liable for public health, extra costs firms and governments bring about to shield supply chains from counterfeit products, resulting in disincentives to foreign investment, and resulting loss of occupations and monetary chances. Unfortunately, the exact extent of this problem, and therefore how best to combat it, is yet obscure.

Sudan is a vast African country surrounded by a number of other African countries (e.g Kynea, Uganda… etc). Most of these African countries suffer from the circulation of such drugs. To combat this problem, these countries have put strategies to control this problem through enforcing strong close and carry out continuous Post Marketing Surveillance (PMS) studies.

The role of pharmaceutical quality control (QC) and Good Manufacturing Practices (GMP) is to assess the quality of active pharmaceutical ingredients (APIs) and excipients [28]. Even though the National Medicine Regulatory Authorities (NMRAs) have been executed in all developing countries, most of them are not completely operational, while the rest at various degrees of foundation.

Many challenges facing authorities, health care providers and National Quality Control Laboratories (NQCL) regarding counterfeit and substandard drugs these include that they are difficult to trace, their spreading can’t be controlled or stopped, their detection, quantification and control need well-equipped labs and well trained trustful personnel and some expired legitimate drugs can be remarked with a false new expiry date. In addition to that counterfeiters aim to avoid raising suspicion about the origin and the quality of their products also they take measures that make them slip past the authorities control and ultimately deceive consumers.

Gaps in current literature include documentation of pharmacist’s knowledge regarding counterfeit medications, strategies used by pharmacists to handle counterfeit medications delivered to their practice setting, and how patients are educated about counterfeit medications. Therefore, this study aimed to reveal the extent of the counterfeit practice in Khartoum locality, Sudan and to assess the public’s experience, attitude and knowledge about CFMs.

## Methods

### Setting and study population

The study was conducted in Khartoum city, Sudan between December 2019 and April 2020. The inclusion criteria were any individual 18 years old and above, willing to participate in the study.

### Sampling procedure

The sample size for public (respondents/ participants) was calculated using the following equation [29]. Participants were then selected randomly.

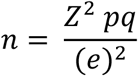

Where: n: sample size; Z: a value from the normal distribution related to 0.05 precision; p: proportion of interest; q: 1-p; e: precision level (0.05).

### Questionnaire design and validation

The questionnaire used different types of questions: multiple-choice, close ended, and open-ended (Supplementary file 1). It consisted of general information and demographic, medicine use, awareness of CFM, attitude towards CFM and recommendations on measures to be taken to control the illicit trade in counterfeit drugs.

The questionnaire was piloted on 20 members of the public for content validation. It was then simplified, shortened to consume 12-15 minutes. Finally, the questionnaire was assessed through experts in the field of Pharmacy before data collection.

### Data collection

The questionnaire was initially written in English, and was translated into Arabic. Authors and assistants (surveyors) were then administered the questionnaire through face-to-face interview for public. The collected data were checked for completeness, manually scored and finally coded before the analysis. The total awareness scores were 10; therefore, those with a total score (1-3) were classified as having a low level of awareness, (4-7) as moderate level, and (8-10) were considered having a high level of awareness. Concerning the attitude, the total score was 16. Those with a total score (1-5) were classified as having a poor attitude, [6–11] as fair attitude, and (12-16) were considered having good attitude.

### Statistical analysis

Data were entered and analysed using Excel 2016. Descriptive analysis was performed for the questionnaire where frequencies, percentages and graphical representation were reported for all categorical variables. Fisher exact test and Spearman correlation was then used for testing the association between variables and the relation of awareness to attitude, respectively. *P* < 0.05 was considered statistically significant.

### Ethical requirements

The ethical clearance (FPEC-03-2019) was obtained from the Ethical Committee of the Faculty of Pharmacy, University of Khartoum. The dates and times when the questionnaires were administered were all documented.

With respect to respondents’ autonomy and anonymity, all of them signed a written informed consent and were guaranteed privacy and confidentiality.

## Results

### Public awareness and attitude

#### Demographic characteristics

A total of 386 respondents were participated in this study; the mean age was 34 years (18-80 years old). About 218 (56%) were males and 194 (50%) were university graduates. Obtained data are summarized in Table 1.

**Table 1:** Socio-demographic Characteristics of the participants (n= 386)

|  | N | (%) |
| --- | --- | --- |
| **Gender** |  |  |
| Male | 218 | (56%) |
| Female | 160 | (41%) |
| Missed | 8 | (3%) |
| **Education Level** |  |  |
| Illiterate | 8 | (2%) |
| Primary school | 18 | (5%) |
| High secondary school | 81 | (21%) |
| University | 194 | (50%) |
| Postgraduate level | 77 | (20%) |
| Missed | 8 | (2%) |
| **Employment Status** |  |  |
| Student | 71 | (18%) |
| Trader | 52 | (3%) |
| Labour | 52 | (13%) |
| Unemployed | 50 | (13%) |
| Housewife | 47 | (12%) |
| Employee | 50 | (13%) |
| Doctor | 11 | (3%) |
| Banker | 8 | (2%) |
| Driver | 5 | (1%) |
| Manager | 5 | (1%) |
| Accountant | 3 | (1%) |
| Teacher | 2 | (1%) |
| Other | 22 | (6%) |
| Missed | 8 | (3%) |
| **Profession** |  |  |
| Non-medical field | 283 | (73%) |
| Medical field | 89 | (23%) |
| Missed | 16 | (4%) |

### Awareness about counterfeit medicines

Most of the respondents 222 (58%) had heard the term “Counterfeit Medicine” before, while 151 (39%) did not know. Social media was mentioned by the majority of the respondents (112) as the main source of awareness, followed by pharmacy (68), TV (37) and billboards (4). Regarding to CFM quality, 73% considered it with worse quality, 22% of same quality and 5% of better quality.

Analysis of questions on how respondents dealt with suspected CFMs showed that 57 (15%) of the respondents were reported being suspicious of different medicines to be counterfeited, such as insulin injections and atorvastatin. Their reasons behind being suspicious and their action towards that are summarized in Table 2.

**Table 2:**
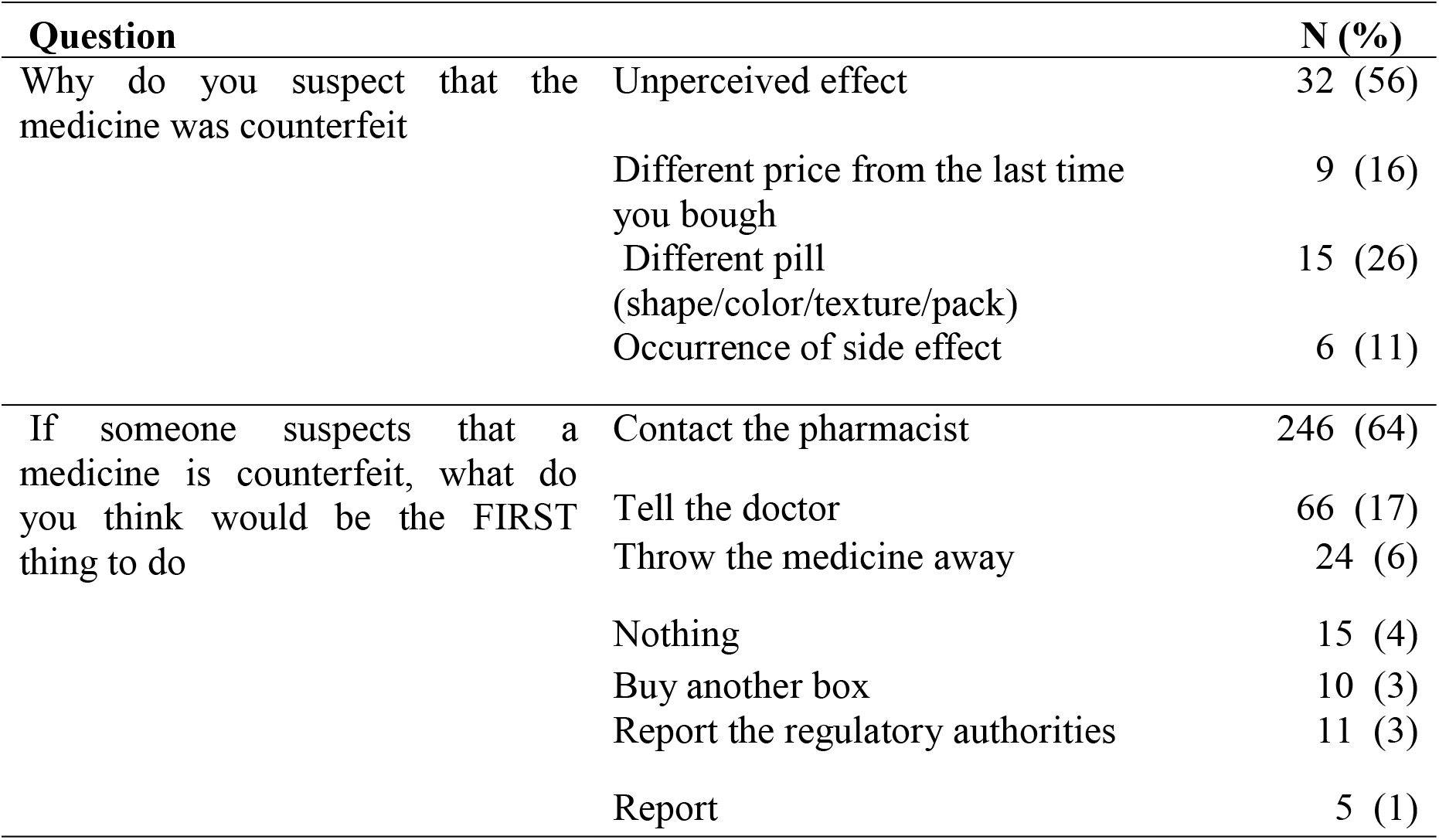
Reason and action toward suspected medicines.

| Question |  | N (%) |
| --- | --- | --- |
| Why do you suspect that the medicine was counterfeit | Unperceived effect | 32 (56) |
|  | Different price from the last time you bough | 9 (16) |
|  | Different pill (shape/color/texture/pack) | 15 (26) |
|  | Occurrence of side effect | 6 (11) |
| If someone suspects that a medicine is counterfeit, what do you think would be the FIRST thing to do | Contact the pharmacist | 246 (64) |
|  | Tell the doctor | 66 (17) |
|  | Throw the medicine away | 24 (6) |
|  | Nothing | 15 (4) |
|  | Buy another box | 10 (3) |
|  | Report the regulatory authorities | 11 (3) |
|  | Report | 5 (1) |

The respondents were further asked about how to avoid buying CFMs, 271 of them (70%, N= 473) answered by getting the medicine from a trustworthy pharmacist while 66 reported buying only medicines manufactured outside Sudan

### CFMs characteristics, origin and availability

The respondents reported different characteristics to distinguish the CFMs. A total response of 642 was obtained. 56% of them mentioned the side effects as main characteristic to identify the CFMs. 52% mentioned those with less effect followed by less price (32%) and different package (30%).

In regards to the source of CFMs, 229 (66%) of the respondents reported Egypt as the main origin of CFMs. Also, when the respondents were asked about whom is responsible for CFMs’ availability in Sudan, 329 (85%) mentioned manufactures followed by companies 259 (67%) (Table 3).

**Table 3.** CFMs source and who is responsible for availability.

| Question |  | N | % * |
| --- | --- | --- | --- |
| In your opinion, from which country do you think most counterfeit medicines originate | Egypt | 229 | 66% |
|  | India | 50 | 14% |
|  | KSA | 8 | 2% |
|  | USA | 7 | 2% |
|  | Chad | 13 | 4% |
|  | Nigeria | 4 | 1% |
|  | Pakistan | 6 | 2% |
|  | Other (South Africa, third world countries, Israel...) | 110 | 32% |
| In your opinion, who is responsible for the availability of counterfeit medicines | Manufacturers | 329 | 85% |
|  | Companies | 259 | 67% |
|  | Pharmacists | 254 | 66% |
|  | Customs | 233 | 61% |

\* Total is more than 100% as more than one answer was possible

### Measures to Reduce Counterfeit Medicines

Most respondents, 316 (82%), reported that counterfeit drugs would be successfully combated through education and sensitizing the public’s awareness (83%) against counterfeit medicines. 312 of the respondents (81% of the total respone) reported campaigns conducted by the Ministry of health for enriching their awareness, 282 (73%) through social media while 229 (59%) believe that the better way to be educated from their pharmacists through brochures (Table 4). The respondents also reported punishment (155 (40%)) and regulation of medicines entry (180 (54%)) as a major role of the regulatory authorities to combat the CFMs problem.

**Table 4.**
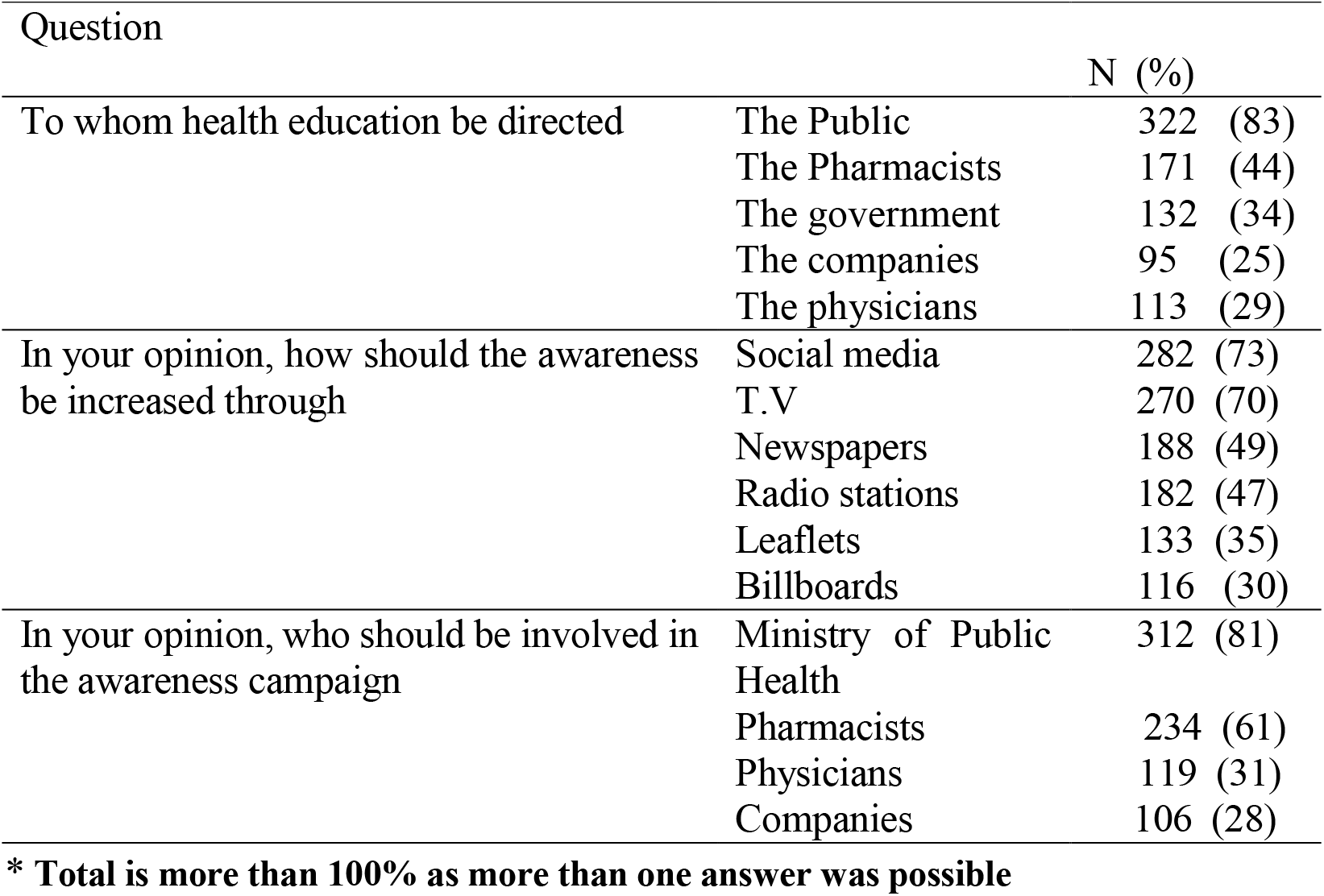
Awareness source and campaigns.

| Question |  | N (%) |  |
| --- | --- | --- | --- |
| To whom health education be directed | The Public | 322 | (83) |
|  | The Pharmacists | 171 | (44) |
|  | The government | 132 | (34) |
|  | The companies | 95 | (25) |
|  | The physicians | 113 | (29) |
| In your opinion, how should the awareness be increased through | Social media | 282 | (73) |
|  | T.V | 270 | (70) |
|  | Newspapers | 188 | (49) |
|  | Radio stations | 182 | (47) |
|  | Leaflets | 133 | (35) |
|  | Billboards | 116 | (30) |
| In your opinion, who should be involved in the awareness campaign | Ministry of Public Health | 312 | (81) |
|  | Pharmacists | 234 | (61) |
|  | Physicians | 119 | (31) |
|  | Companies | 106 | (28) |

\* Total is more than 100% as more than one answer was possible

Using the scoring system, the obtained results indicated a fair level of awareness in 80% of the respondents, while the good and poor levels of awareness were represented by 2% and 18% of the respondents, respectively.

### Public views towards CFMs

Public attitude was assessed using statements describing the quality, risk, experiences, and CFMs’ price. Obtained data are shown in Figure 1 as agree, neutral and disagree based on the respondents’ view.

**Figure 1:**
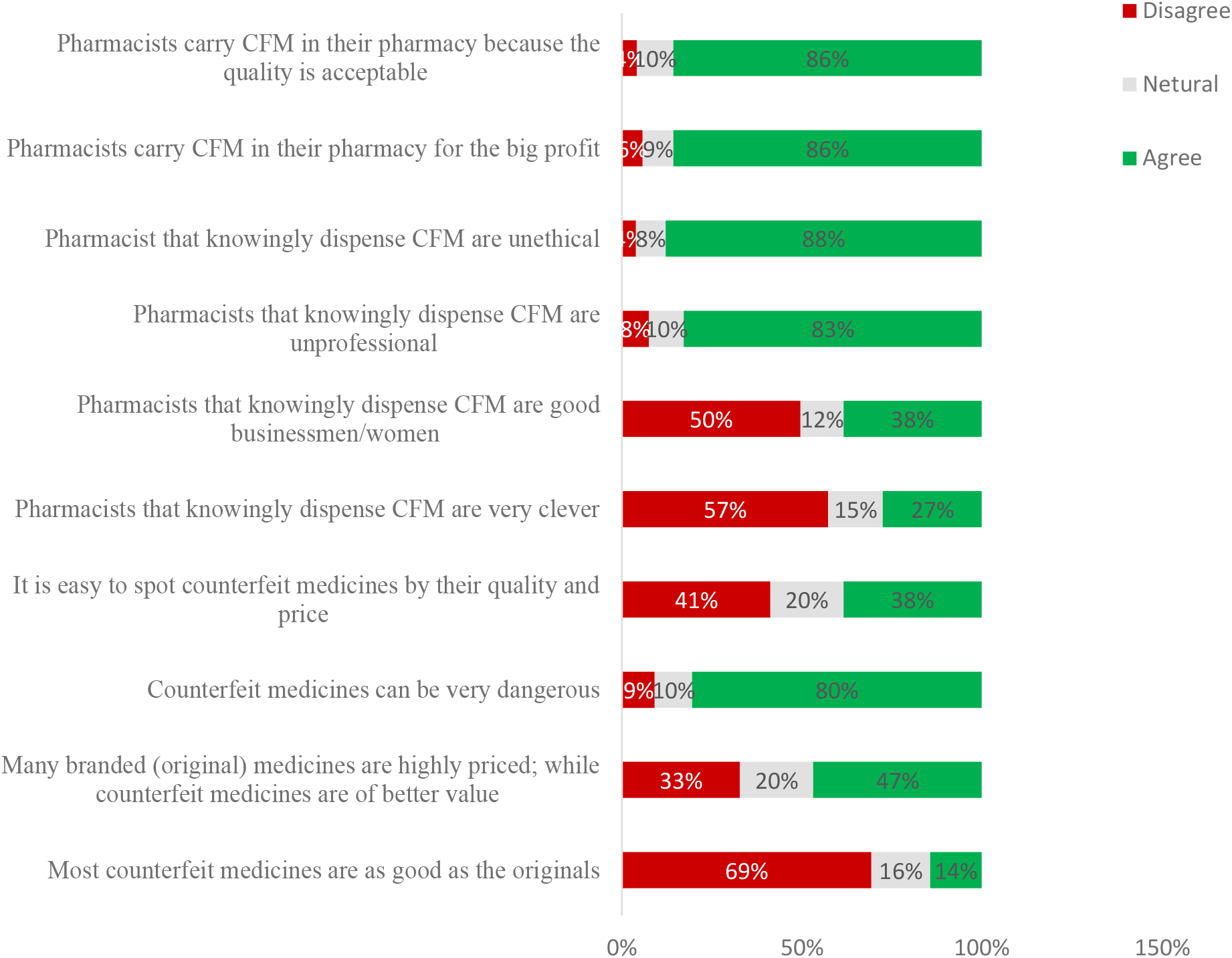
Respondents’ views and experience towards CFMs (N=384)

Regarding the overall attitude, a good attitude was found in 68% of the respondents, while poor and fair attitude were shown in 2% and 30% of the respondents, respectively. Fisher exact test revealed no significant relation to the tested variables.

Fisher exact test revealed a significant association between the attitude, gender and TV (P value = 0·011 and 0·003, respectively), while only social media was found to be significantly related to awareness (P value = 0·002). However, socio-demographic characteristics, profession and education level were not borne out to be significantly associated with the awareness and attitude.

Spearman correlation is a bivariate correlation measure suitable for the analysis of data that is not normally distributed. The correlation is measured between −100% to 100%, the negative indicates a negative relationship that is as one increases the other decreases. This test was thus applied to assess the correlation between awareness and attitude. A positive correlation of 16.9% was obtained with statistical significance (P value = 0·001) indicating increasing the awareness will have a positive effect on the attitude.

## Discussion

Counterfeit medicines are deadly and growing problem. According to WHO, as many as one in ten medical products circulating in developing countries is substandard or falsified. The problem is particularly common within Africa. Of all the reported fake drugs to the WHO between 2013 and 2017, 42% of them came from African region. In 2019, WHO raised alerts for this problem in Niger, Cameron, Kenya and Uganda [13]. Food and Drug Administration (FDA) has also developed a reporting system for suspicion about CFMs through med watch [30]. Although the issue is widely acknowledged, its complexities are poorly understood especially in Africa as no official data is available.

Clearly, there is work to be done from a structural perspective. The WHO is working with the African Union to improve health coverage across Africa. Governments must use the available technology to create more visible supply chains.

To best of our knowledge, this is the first study to evaluate the CFMs in Sudan. The public’s awareness, experience and view towards CFMs have been explored in Sudan and assessed in order to come up with substantial measures and recommendation to help combating the widely spread CFMs issue.

In our present study, the mean age of participants was found to be 34years old, with the acute condition indication had been reported by high percentage of respondents rather than chronic indication. Different education levels were existed in the sample with the highest percentage for the university level.

The majority of the participants reported that they prefer to buy their medicines from the same pharmacy due to knowledgeable pharmacists and medicines’ availability. It was also indicated that restriction to certain brand name is attributed to high perceived effect for medicines.

In regards to the awareness about CFMs, 222 (58%) of the respondents reported that they are aware of the term compared to 93.4% reported in Sholy’s study [31]. Social media was the most mentioned source for the awareness in contrast to T.V in Lebanon study, considering Egypt and India being the main origin for this problem.

WHO provided information for consumers when suspected a medicine is counterfeited to first, talk to the pharmacist whom you bought the medicine from or contact the healthcare professional for medical advice [32]. FDA voluntary reporting system named Med watch is also available for consumers and health care professionals [30]. In our study, about three quarter of the respondents reported that the quality of CFMs is less than the authentic ones, which is expected, since CFMs are defined to contain wrong or less active pharmaceutical ingredient than the stated. Different classes of medications are mentioned as a case of suspicion due to unperceived effect being mentioned as the most reason. Although some of the respondents did nothing or throw the medicine indicating that they are not educated from responsible authority on how to deal with suspicion, the majority of them responded in accordance to the WHO guides by either contacting the pharmacists or telling the doctor (Figure 3). The recognition of counterfeit/fake drugs by the public is thus expected to have a great impact on decreasing the purchase of such drugs in the market.

A substantial number of respondents reported to distinguish between genuine and CFMs, and buying their medicines from trustworthy pharmacists in order to protect themselves. Percentage of respondents who chose “causing side effects” and “producing less effect” as characteristics for CFMs were more than with “less price” and “different packaging”, this might be due to their lack of knowledge about the packaging materials and their deep concern with medicine’s effect and risk rather than cost and packaging.

One major contributing factor to the prevalence of counterfeit medicines in a country is the lack of knowledge and awareness of the society [33, 34]. This was agreed by 316 (82 %) of the respondents suggesting that education will have a vital role in combating the problem. Social media and T.V. were established as the main sources of information and education which is similar to the Cotonou study that referred to television, followed by radio, as convincing sources of information [35]. Respondents reported that education should be directed to different parties of the community such as pharmacists, companies and regulatory authorities, and must be carried out by different bodies such as the Ministry of Health through workshops and campaigns conducted at least twice a year.

Moreover, the weak control on chain supply is an additional factor for the spread of CFMs which has been reported by different studies [36, 37]. Respondents believe that manufacturers were responsible for the availability of CFMs.

Thus, counterfeit drugs will be successfully combated through strict regulation and different punishment strategies to handle the offenders (regulation specifically, control of entry port and quality control).

With respect to the attitude of respondents towards pharmacists’ dealing with CFMs, although 38% of them agreed that they are business men/women, the majority believed that they are unprofessional and unethical and dealing with CFMs for the profit and easy money. A high percent of the respondents (86%) agreed that pharmacist carry CFMs in their pharmacies since the quality is acceptable and many branded (original) medicines are highly priced and not available while CFMs are of better value. These results are contrary to their response about the quality of CFMs compared to authentic ones, indicating in this point that respondents’ attitude does not match their awareness. Although different studies reported that people realized that counterfeits were inferior to originals [38–40], their superior prices might compensate for the lower quality and efficacy [41]. This could be the case for the members of the public with low socioeconomic status. Alfadl et al, 2012 examined the influence of certain factors on consumers’ behaviour regarding CFMs in Sudan, and found that motivation and subjective norms were positively and significantly related to purchase intent of CFM, but not attitude [42]. Additionally, the findings suggested that Sudanese consumers might be motivated to buy CFMs when medicines are inaccessible and/or unaffordable, and these were considered the main contributors to buying CFMs in developing countries. Therefore, controlling the cost of medicine needs to be considered, and evaluated by pharmaceutical companies and health authorities.

38% of the respondents believed it is not easy to spot CFMs by the price and quality. Previous studies reported that patients were confused and unsure if “generic brands” of medicines were in fact counterfeits [43]. Many studies also reported that, lack of patients’ knowledge about CFMs led them to have more negative attitudes towards generic medicines [44, 45]. Therefore, education and awareness are important as generics of essential medicines have a global public health benefit, because they are less expensive and more accessible to people [46]. Consequently, according to respondents of the public awareness survey reported that the best way to avoid buying CFM, was going to a trustworthy pharmacist.

Although, there is a positive correlation between awareness and attitude, socio-demographic characteristics, profession and education level were not borne out to be significantly associated with the awareness and attitude. This result is inconsistent with the Mhando et al and Sholy et al results, which they reported that education and profession has a significant effect on awareness and attitude [31, 47].

The limitation of the present study is that the population may not appear diverse enough; as respondents were reported to be mainly from Khartoum city. Additionally, the cross-sectional design of the study not allow generalization of the findings to all community pharmacists in Sudan.Further studies may be needed in other cities and rural areas of Sudan to assess the awareness and attitude towards CFMs. Despite this, our study would be the groundwork for future studies in CFMs as community involvement is a neglected issue in the fight against the illicit trade in counterfeit drugs.

## Conclusion

The present study concluded that most of the respondents were aware about CFMs with positive attitude towards it. Social media was reported as the main source of information by the respondents. No significant correlation between variables was reported. Thus, it is highly needed to conduct educational campaigns, emphasizing the risks and consequences associated with CFMs, in addition to addressing the public demands for CFMs to strengthen laws and regulation and increase public and other health care professionals’ awareness toward CFMs.

## Data Availability

All dataset supporting this article is available within the article and its supplementary files.

## Acknowledgements

Authors acknowledge Faculty of Mathematical sciences, University of Khartoum, Sudan for the sample calculation and statistical analysis. Authors are also thankful for Dr. Bashir Y. For revising the questionnaire and drafts; the surveyors, Mr. Mohamed A., Ms. Roaa E., Mr. Khalid M., Ms. Sara O., Ms. Rana E., Ms. Ayat I., who helped in the data collection and entry.

## Data availability

All dataset supporting this article is available within the article and its supplementary files.

## Author contribution

S.S. wrote the paper draft, edit and revise it, conducted part of data entry; W.W wrote the results and discussion, conducted part of data collection and entry; E.A. revise the final draft.

